# A comprehensive Indian clinical and imaging Coronary Artery Disease Dataset: ICADD

**DOI:** 10.1101/2023.05.13.23289934

**Authors:** Akanksha Bohra, Ashita Barthur, Rahul Patil, Santosh Ansumali

**Affiliations:** Jawaharlal Nehru Centre For Advanced Scientific Research, Jakkur, Bengaluru, Karnataka 560064; Sri Jayadeva Institute of Cardiovascular Sciences and Research, Bengaluru, Karnataka 560041

## Abstract

**Background & objectives:** Coronary artery disease (CAD) is one of the leading causes of mortality worldwide and contributes significantly to the disease burden in India. The dearth of datasets for Indian patients with CAD is an obstacle to further development and collaborative work in clinical research. The purpose of this study was to create a comprehensive dataset of diagnostic information on CAD in Indian patients. We present Indian Coronary Artery Disease Dataset (ICADD) with CT and invasive FFR data to aid innovation and improvement of non-invasive diagnostic procedures.

**Methods:** The data was collected in a tertiary care centre in South India. The patients with suspected CAD who underwent coronary CT-angiography (CCTA) and invasive angiography (ICA) with the measurement of fractional flow reserve (FFR) were included in the study.

**Results:** The data from 70 patients were collected between April 2018 and Dec 2021. The dataset contains 70 CCTA, 70 ICA scans along with FFR, and echocardiography and haematology reports. It includes 54 (77.14%) male and 16 (22.85%) female patients. The mean age of participants is 50.7±8.8years.

**Conclusions:** To the best of our knowledge, ICADD is the first of its kind database in India. This dataset can be used to create a benchmark database, serve as reference for performance and evaluation of clinical utility of novel non-invasive diagnostic methods and additionally serve as a database for clinical research.

## Introduction

According to world health organisation (WHO) statistics, cardiovascular diseases caused 17.9 million deaths (32% of total deaths globally) in 2019^**1**^. Among all cardiovascular diseases, coronary artery disease (CAD) alone was the reason for 8.9 million deaths (16 % of total deaths) in 2019^**2**^. In the Indian context, cardiovascular diseases contributed to 28.1% of total deaths in 2016 with CAD alone causing 17.8% of total deaths^**3**^. Current management for CAD involves diagnosis using one of the imaging modalities followed by percutaneous interventions or surgery, and or medications. Early detection and accurate diagnosis can be highly effective in reducing mortality rate from CAD^**4**^. Diagnostic methods include Electrocardiography (EKG), Echocardiography (ECHO), coronary computed tomography-angiography scan (CCTA), and magnetic resonance imaging (MRI), single photon emission computed tomography (SPECT), PET (Positron emission tomography) and conventional or invasive coronary angiography (ICA) are used in case of CAD. Typically, CCTA has a high negative predictive value and is the established diagnostic method to detect CAD^**5**^. Invasive fractional flow reserve (FFR) measurement along with ICA allows combined anatomic and functional evaluation of the disease. Invasive FFR is considered a ‘gold standard’ for severity assessment of vessel-specific ischaemia because of its accuracy^**6**^. The invasive FFR is a hyperaemic index. There are non-hyperaemic indices like instantaneous wave-free ratio (iFR) and resting full-cycle ratio (RFR) which have been shown to be non-inferior to invasive FFR ^**7, 8, 9**^. To minimize the risks, complications and costs associated with invasive procedures, there is recent emergence in non-invasive methods for early diagnosis of CAD through machine learning and computational fluid dynamics modelling ^**10, 11, 12**^. Coronary CT-scan derived FFR (CT-FFR) is a non-invasive method for functional assessment of the disease which has been clinically validated in multicenter trials ^**13,14, 15**^.

Even though various heart disease and CT scan databases are available ^**16, 17, 18**^, none of existing datasets include invasive FFR information or address the importance of diagnostic information obtained through multiple methods (CCTA, ICA, invasive FFR or invasive RFR) for the same patient. Such datasets consisting of data of Indian patients are even more scarce. Therefore, we present the Indian Coronary Artery Disease Dataset (ICADD). The purpose of this study was to create a comprehensive dataset of diagnostic information of CAD in Indian patients with invasive FFR data to aid innovation and improvement of non-invasive diagnostic procedures. In particular, each dataset in ICADD includes CCTA data with corresponding functional index of invasive FFR or invasive RFR for intermediate stenosis. The dataset also features other fundamental information for diagnosis of CAD such as demographic and comorbidities information, haematology reports and the data on associated cost of these clinical procedures. This integrated dataset can be used as an objective benchmark to evaluate effectiveness of CT-FFR for Indian patients and for development of new algorithms and automated classifications models for non-invasive diagnosis in addition to acting as a pilot for other clinical research.

## Methods

The data was collected in a tertiary care centre in South India, between Apr 2018 and Dec 2021. All the subjects were patients with suspected CAD who underwent CCTA, ICA with the measurement of fractional flow reserve (FFR). The study was approved by the institutional ethics committee. CCTA and FFR were performed as a part of routine clinical care. Consenting adults 18 years of age or older, for whom all three datasets, including CCTA, ICA and FFR or RFR were available were included in the study. We collected the demographic, clinical, lab and ECHO details of the patients, when available. All the collected datasets were de-identified and each patient was assigned with an unique study-id.

### Coronary CT-scan

CCTA was performed on a Brilliance 64-slice CT scanner (Philips, Best, The Netherlands) with an aperture of 70cm. Patients were screened for contraindication to β -blocker and nitro-glycerine (NTG). Patients were also screened for health conditions like renal insufficiency and previous heart failure. CCTA was performed on patients with serum creatinine <1.5mg/dl or glomerular filtration rate(GFR) >45 mL/min and heart rate <65 bpm according to institution protocol. The intravenous beta-blocker metoprolol was administered targeting a heart rate of <65 beats/min in patients with higher heart rates. Immediately before image acquisitions, nitro-glycerine (NTG) was administered to ensure coronary vasodilation. During acquisition, iodine contrast was injected which was followed by a saline flush. CCTA were obtained in helical mode with retrospective electrocardiographic gating with tube voltage of 120mV. The spatial resolution of a CT image acquired at is 24-line pairs/cm and 512x512 pixels with 0.5 mm slice spacing and 0.4x0.4mm pixel spacing.

### Invasive Coronary Angiography

ICA procedure was performed with the Allura Xper FD10 Cath/Angio System (Philips, Best, The Netherlands). Standard protocol was followed for ICA. It was obtained with three standard views, RAO caudal, RAO cranial, LAO caudal and one additional projection for the optimum view of the stenosis.

### Invasive FFR

Invasive FFR was performed at the time of ICA for clinically indicated vessels. The invasive FFR value was measured using 0.014inch PressureWire Aeris with Agile Tip (St. Jude Medical Systems AB (Abbott) through femoral or radial approach. The calibration and pressure equalization were performed according to guidelines. Nitroglycerin was administered to minimize vessel resistance before measuring FFR. The intravenous adenosine was administered to induce hyperaemia for FFR measurement. The RFR measurements were made without adenosine. The FFR value of ≤0.8 and the RFR value of ≤0.89 for stenosis was considered ischaemic. The revascularization decisions were taken according to standard practices by the performing interventional cardiologist.

## Results

The ICADD dataset contains the data of 70 patients. The available data is listed in Table 1. For each patient, it includes the corresponding CCTA and ICA data along with echocardiogram and haematology reports wherever available, thus including a total of 70 CCTA data, 70 ICA data and 53 echocardiogram reports (Table 1). Baseline characteristics of study population are tabulated in Table 2. Categorical data are described using frequencies and percentage and continuous data using mean and standard deviation. Total 70 patients met the inclusion criteria. 54 (77.14%) were males and 16 (22.86%) were females. The mean age of the subjects was 50.7 years (range from 23 to 68). The mean time interval between CCTA and ICA procedure was 17.7 days and standard deviation was ±30.4 days. The details on cardiovascular risk factors were available for 69 subjects. The main risk factors were hypertension (57.9%), diabetes (40.5%), smoking (17.4%), alcoholism (5.8%) and dyslipidaemia (4.3%) in the study population.

**Table 1:** Summary of the data set

|  |  |
| --- | --- |
| Total no. of patients | 70 |
| Total no. of coronary CT-scans | 70 |
| Total no. of ICA scans | 70 |
| Total no. of Haematology reports | 69 |
| Total no. of Echocardiogram reports | 53 |
| No. of patients with invasive FFR measurement | 29 |
| No. of patients with invasive RFR measurement | 30 |
| No. of patient with both FFR and RFR measurement | 11 |

**Table 2:**
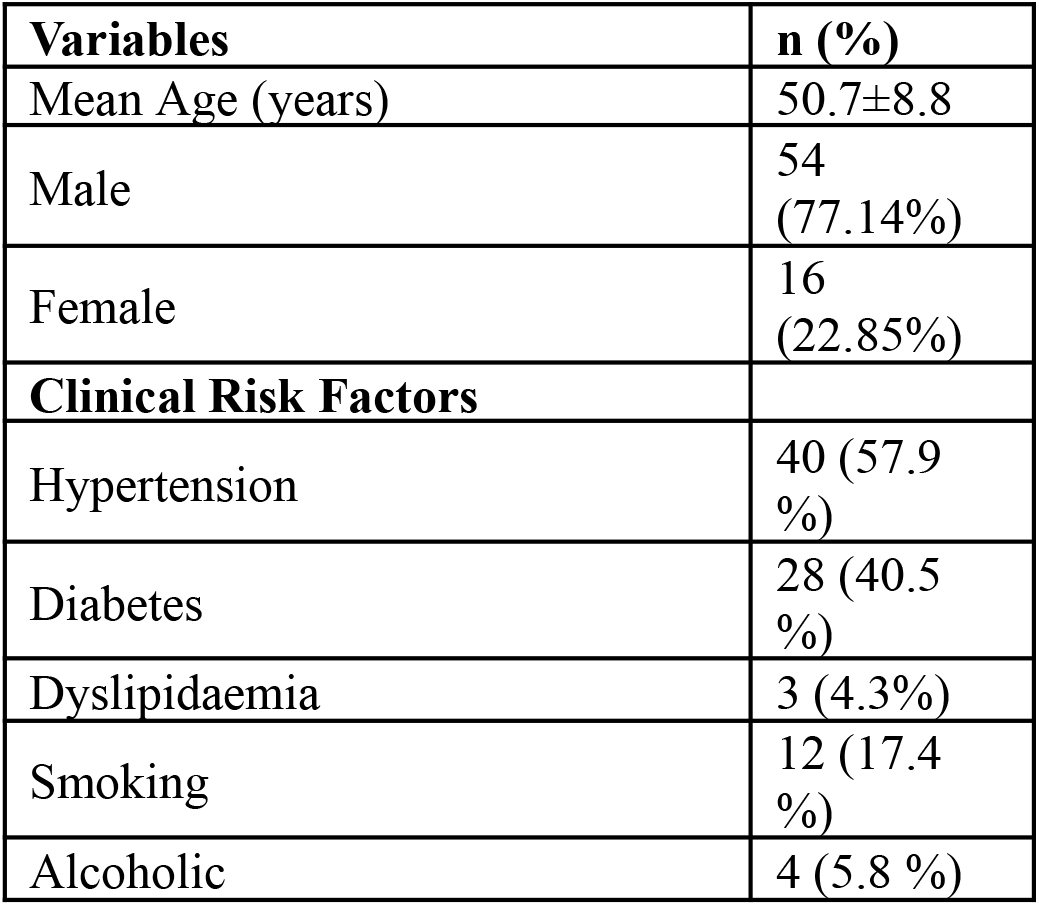
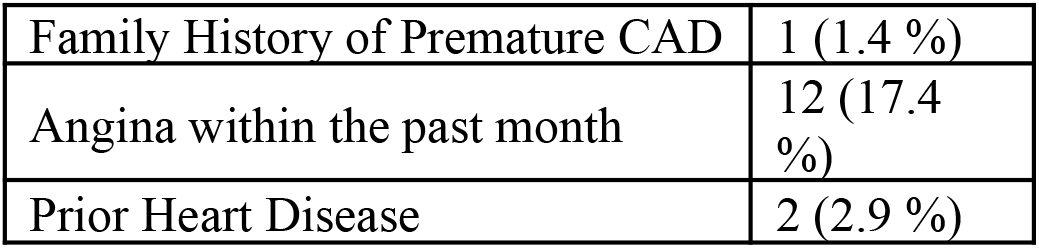
Baseline characteristics of study population

The database also includes haematology reports for most of the patients, however the availability of parameters varies across patients. The mean values for some of the data points is listed in Table 3 with respective sample size available. Lipid profile data was available for 20 patients. The invasive FFR values are available for 29 patients and invasive RFR are available for 30 patients. Both values of invasive FFR and RFR are available for 11 patients. The FFR and RFR are recorded with the time-series data during the measurements. The 2D-echocardiographic reports include measurements such as left ventricular systolic and diastolic dimensions, end-diastolic and end-systolic volumes, ventricular wall thickness and ejection fraction.

**Table 3:**
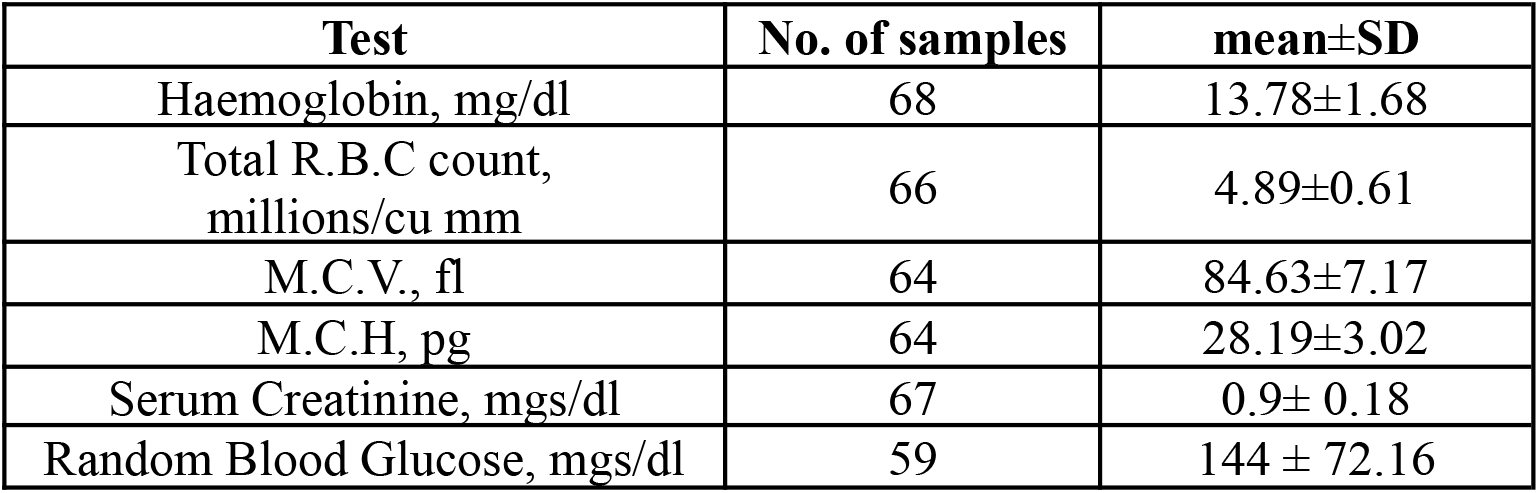
Haematology Parameters

## Discussion

The primary aim of this study was to create a comprehensive data set which included CCCTA images, ICA images and FFR measurements for Indian patients. To the best of our knowledge, this is the first comprehensive database of Indian patients with the availability of supplementary clinical information.

Most of the datasets available on the internet are curated towards machine learning and deep learning methods and only contain tens of images. Some of the datasets provide a full DICOM set^**19**^ but include only either CT-scan or invasive angiography data. We present “real world” data which contains the complete set of DICOM images with original resolution for both the CCTA and ICA. The data has not been labelled or annotated. The dataset can be used to create a benchmark database and serve as reference for future collaborative work. This dataset adds to the already available larger data sets and will be a valuable resource to further research on Indian subset. The data can be used to establish the performance of non-invasive diagnostic methods like CT-FFR. It also provides insight into anatomical and disease patterns and prevalence.

## Data Availability

All data produced in the present study are available upon reasonable request to the authors

## Availability of Data

The datasets acquired during the current study are available from the corresponding author on request. It may require a sub-contract for data access or service fee.

## Notes

### Competing Interest Statement

The authors have declared no competing interest.

### Funding Statement

The study was funded through a grant provided by Rajiv Gandhi University of Health Sciences, Bengaluru for a collaborative project (Project ID: 17C018B) between Sri Jayadeva Institute of Cardiovascular Sciences and Research, Bengaluru and Jawaharlal Nehru Center for Advanced Scientific Research, Bengaluru.

### Author Declarations

Institute ethics committee of Sri Jayadeva Institute of Cardiovascular Sciences and Research, Bengaluru, India 560041 gave ethical approval for this work.

## References

1. World Health Organization, Cardiovascular diseases (CVDs) [page on the internet].[updated 2021 June 11]. Available from: https://www.who.int/en/news-room/fact-sheets/detail/cardiovascular-diseases-(cvds), accessed on December 01, 2022.

2. World Health Organization, The top 10 causes of death [updated 2020 December 9]. Available from: The top 10 causes of death, https://www.who.int/news-room/fact-sheets/detail/the-top-10-causes-of-death), accessed on December 01, 2022.

3. The changing patterns of cardiovascular diseases and their risk factors in the states of India: the Global Burden of Disease Study 1990–2016, Lancet Glob Health 2018; 6: e1339–51, Published Online September 12, 2018. Available from: http://dx.doi.org/10.1016/S2214-109X(18)30407-8, accessed on October 02, 2022.

4. Arnett DK, Blumenthal RS, Albert MA, Buroker AB, Goldberger ZD, Hahn EJ, Himmelfarb CD, Khera A, Lloyd-Jones D, McEvoy JW, Michos ED, Miedema MD, Muñoz D, Smith SC Jr, Virani SS, Williams KA Sr, Yeboah J, Ziaeian B. 2019 ACC/AHA Guideline on the Primary Prevention of Cardiovascular Disease: A Report of the American College of Cardiology/American Heart Association Task Force on Clinical Practice Guidelines. Circulation. 2019 Sep 10;140(11):e596–e646. doi: 10.1161/CIR.0000000000000678. Epub 2019 Mar 17. Erratum in: Circulation. 2019 Sep 10;140(11):e649-e650. Erratum in: Circulation. 2020 Jan 28;141(4):e60. Erratum in: Circulation. 2020 Apr 21;141(16):e774. PMID: 30879355; PMCID: PMC7734661.

5. Budoff MJ, Dowe D, Jollis JG, et al. Diagnostic performance of 64-multidetector row coronary computed tomographic angiography for evaluation of coronary artery stenosis in individuals without known coronary artery disease: results from the prospective multicenter ACCURACY (Assessment by Coronary Computed Tomographic Angiography of Individuals Undergoing Invasive Coronary Angiography) trial. J Am Coll Cardiol 2008;52:1724–32.

6. Zimmermann, F. M., Omerovic, E., Fournier, S., Kelbæk, H., Johnson, N. P., Rothenbühler, M., et al. 2019 Fractional flow reserve-guided percutaneous coronary intervention vs. medical therapy for patients with stable coronary lesions: Meta-analysis of individual patient data. European Heart Journal 40 (2), 180–186.

7. Berry C, van ‘t Veer M, Witt N, Kala P, Bocek O, Pyxaras SA, et al. VERIFY (VERification of Instantaneous Wave-Free Ratio and Fractional Flow Reserve for the Assessment of Coronary Artery Stenosis Severity in EverydaY Practice): a multicenter study in consecutive patients. J Am Coll Cardiol. 2013 Apr 2;61(13):1421–7. doi: 10.1016/j.jacc.2012.09.065. Epub 2013 Feb 6. PMID: 23395076.

8. Götberg M, Christiansen EH, Gudmundsdottir IJ, Sandhall L, Danielewicz M, Jakobsen L, et al. Instantaneous Wave-free Ratio versus Fractional Flow Reserve to Guide PCI. N Engl J Med. 2017 May 11;376(19):1813–1823. doi: 10.1056/NEJMoa1616540. Epub 2017 Mar 18. PMID: 28317438.

9. Svanerud J, Ahn JM, Jeremias A, van ‘t Veer M, Gore A, Maehara A, et al. Validation of a novel non-hyperaemic index of coronary artery stenosis severity: the Resting Full-cycle Ratio (VALIDATE RFR) study. EuroIntervention. 2018 Sep 20;14(7):806–814. doi: 10.4244/EIJ-D-18-00342. PMID: 29790478.

10. Tesche C, De Cecco CN, Baumann S, Renker M, McLaurin TW, Duguay TM, et al. Coronary CT Angiography-derived Fractional Flow Reserve: Machine Learning Algorithm versus Computational Fluid Dynamics Modeling. Radiology. 2018 Jul;288(1):64–72. doi: 10.1148/radiol.2018171291. Epub 2018 Apr 10. PMID: 29634438.

11. Stuckey TD, Gammon RS, Goswami R, Depta JP, Steuter JA, Meine FJ 3rd, et al. Cardiac Phase Space Tomography: A novel method of assessing coronary artery disease utilizing machine learning. PLoS One. 2018 Aug 8;13(8):e0198603. doi: 10.1371/journal.pone.0198603. PMID: 30089110; PMCID: PMC6082503.

12. Alizadehsani R, Abdar M, Roshanzamir M, Khosravi A, Kebria PM, Khozeimeh F, Nahavandi S, Sarrafzadegan N, Acharya UR. Machine learning-based coronary artery disease diagnosis: A comprehensive review. Comput Biol Med. 2019 Aug;111:103346. doi: 10.1016/j.compbiomed.2019.103346. PMID: 31288140.

13. Nakanishi R, Budoff MJ. Noninvasive FFR derived from coronary CT angiography in the management of coronary artery disease: technology and clinical update. Vasc Health Risk Manag. 2016 Jun 22;12:269–78. doi: 10.2147/VHRM.S79632. PMID: 27382296; PMCID: PMC4922813.

14. Curzen NP, Nolan J, Zaman AG, Nørgaard BL, Rajani R. Does the Routine Availability of CT-Derived FFR Influence Management of Patients With Stable Chest Pain Compared to CT Angiography Alone?: The FFRCT RIPCORD Study. JACC Cardiovasc Imaging. 2016 Oct;9(10):1188–1194. doi: 10.1016/j.jcmg.2015.12.026. PMID: 27568119.

15. Nørgaard BL, Gaur S, Fairbairn TA, Douglas PS, Jensen JM, Patel MR, et al. Prognostic value of coronary computed tomography angiographic derived fractional flow reserve: a systematic review and meta-analysis. Heart. 2022 Feb;108(3):194–202. doi: 10.1136/heartjnl-2021-319773. Epub 2021 Oct 22. PMID: 34686567; PMCID: PMC8762006.

16. Dua, D. and Graff, C. UCI Machine Learning Repository [Internet]. UCI Machine Learning Repository; 2019. Irvine, CA: University of California, School of Information and Computer Science. Available from: https://archive.ics.uci.edu/ml/index.php, accessed on November 30, 2022

17. Masoudi M, Pourreza HR, Saadatmand-Tarzjan M, Eftekhari N, Zargar FS, Rad MP. A new dataset of computed-tomography angiography images for computer-aided detection of pulmonary embolism. Sci Data. 2018 Sep 4;5:180180. doi: 10.1038/sdata.2018.180. PMID: 30179235; PMCID: PMC6122162.

18. Manu Siddhartha. Heart Disease Dataset (Comprehensive) [Internet]. IEEE Dataport; 2020. Available from : https://dx.doi.org/10.21227/dz4t-cm36, accessed on November 22, 2022.

19. Segmed [homepage on the Internet]. Available from: https://www.segmed.ai/, accessed on November 22, 2022.

